# Socio-demographic variation in adherence to The Eatwell Guide within the UK Biobank prospective cohort study

**DOI:** 10.1101/2025.06.06.25329110

**Authors:** Alex Griffiths, Fiona Malcomson, Jamie Matu, Sarah Gregory, Andrea Mary Fairley, Rebecca Fay Townsend, Amy Jennings, Nicola Ann Ward, Louisa Ells, Emma Stevenson, Oliver M Shannon

## Abstract

The Eatwell Guide depicts the UK Government’s healthy eating recommendations and is widely used in clinical practice and public health settings. There is limited evidence on whether adherence to the Eatwell Guide differs by socio-demographic characteristics. This study aimed to explore patterns of Eatwell Guide adherence across socio-demographic groups in the UK Biobank cohort. Eatwell Guide adherence scores were derived for 192,825 individuals from 24-hour dietary recall data (Oxford WebQ), and quantified using a graded, food-based scoring system. Eatwell Guide scores were compared between different age, sex, BMI, ethnicity, socioeconomic status and education groups. Data were analysed using independent sample t-tests, and one-way ANOVA with Tukey post-hoc tests. Eatwell Guide adherence was higher for older than younger, and female compared with male participants (both *p*<0.001). There was a main effect of BMI on total adherence (*p*<0.001), with the highest scores achieved by those with a healthy BMI. Eatwell Guide adherence was higher in white vs non-white participants (*p*<0.001), and differed significantly by education level (*p*<0.001), with the highest score achieved by participants with a higher education level. Total adherence scores differed by socio-economic status (all *p*<0.001), with the highest score achieved by the least deprived participants and the lowest score achieved by the most deprived participants. These data demonstrate that Eatwell Guide adherence differs significantly between socio-demographic groups in the UK Biobank. Exploring the consistency of these findings in other cohorts and developing strategies to increase adherence to the Eatwell Guide in groups with low adherence, are future research priorities.

## INTRODUCTION

Consumption of a sub-optimal diet has been associated with a multitude of non-communicable diseases such as obesity (Romieu et al., 2017), cardiovascular disease (Bowen et al., 2018), type II diabetes (Hu et al., 2001), cancer (Key et al., 2004) and dementia (Morris, 2016). These conditions are associated with a significant health and economic burden, with an estimated US$47 trillion (∼£36 trillion) in costs expected to be incurred by the global economy between 2010-2030 due to non-communicable diseases (Bloom et al., 2012).

Disease prevalence is not uniform across all population groups which may, at least in part, be related to different levels of adherence to healthy lifestyle behaviours such as consumption of a healthy diet (Bennett et al., 2018; Malcomson et al., 2023; Carrasco-Marin et al., 2024). For example, Bennett et al. (2018) found that female participants were more likely to exceed recommended intakes of total sugar, total fat and saturated fat, and consume below the recommended intake of dietary fibre when compared to males. In contrast, male participants were less likely to meet recommended intakes of carbohydrates, protein and polyunsaturated fat (Bennett et al., 2018). Malcomson et al. (2023) evaluated the influence of a much broader group of socio-demographic factors on the intake of whole foods (as the dietary component of Cancer Prevention Recommendations), rather than individual nutrients. Their data suggests that younger, male participants, and those with lower educational attainment were less likely to reach fruit and vegetable intake recommendations. In addition, younger, male participants consumed more red and processed meat than their older and female counterparts respectively. Dietary fibre intake was lower than recommendations for all socio-demographic groups, and differences were observed between all socio-demographic groups with the exception of ethnicity.

In the United Kingdom (UK), government healthy eating recommendations are depicted in the Eatwell Guide (PHE, 2016). This provides information on the types and proportions of different foods and drinks an individual should consume to have a healthy, balanced diet. The Eatwell Guide has a particular focus on whole foods, and endorses a dietary pattern characterised by consumption of a variety of fruits and vegetables, moderate consumption of starchy carbohydrates and wholegrains, moderate consumption of beans and pulses, meat, fish and eggs, adequate fluid intake, and low intake of foods rich in fat, salt and sugar (PHE, 2016; Scarborough et al., 2016). Exploring how overall adherence to the Eatwell Guide differs between specific population groups could be beneficial by providing a) a more comprehensive insight into population differences in overall diet quality than focusing on individual nutrients or foods, and b) insight into UK-specific (rather than general) healthy eating recommendations, which may be more relevant to UK public health bodies and the NHS.

In a UK Biobank analysis, Carrasco-Marin et al., (2024) assessed diet quality and associations with socio-economic status using a simplified, binary dietary risk score based on nine food components drawn from UK and European guidelines. They found that individuals from more deprived areas, with lower education and a lower income were more likely to adhere to a poorer diet. Whilst analysis and interpretation of demographic factors was limited, participants with healthier diets were also more likely to be older, female, of white ethnicity, and have a lower BMI. Whilst insightful, these analyses are limited in several respects. Firstly, the dietary score used was not a direct measure of adherence to the Eatwell Guide, but rather a simplified classification of “healthy” versus “unhealthy” dietary patterns based on selected food components. Secondly, the authors used dietary data from the UK Biobank touchscreen questionnaire, which lacks granularity (compared with the more comprehensive Oxford WebQ tool which was also applied in this cohort). As a result, they were unable to quantify intake of key food groups referenced in the Eatwell Guide such as starchy carbohydrates, wholegrains, beans and pulses and discretionary foods, or else used non-quantified proxies (e.g., preference for semi-skimmed milk) for others like dairy. Thirdly, the binary dietary scoring system (assigning 1 or 0 points based on whether guidelines were met) may also fail to capture small variations in intake levels that exist across a population, especially for individuals whose intakes approach, but do not fully meet recommendations.

Against this background, this study aims to describe adherence to the Eatwell Guide and explore patterns of adherence across socio-demographic groups in the UK Biobank cohort. By using a comprehensive, graded food-based Eatwell Guide score we will be able to provide a nuanced evaluation of how adherence to UK-specific healthy eating recommendations differs across sociodemographic groups, with potential to inform public health and policy initiatives.

## METHODS

### Study population and design

This study used cross-sectional data from the UK Biobank, a prospective cohort study of over 500,000 participants evaluating the determinants of disease in the UK population (Ollier et al., 2005). The design and methods of this study have been described previously (Sudlow et al., 2015). Briefly, participants aged 37-73 years were recruited from across England, Scotland and Wales using National Health Service (NHS) patient registers. At the baseline study visit, participants completed a touchscreen questionnaire detailing sociodemographic characteristics, diet, physical activity and general health. Participants also completed a verbal interview and provided measures of physical function alongside biological samples. Additional measures have been added to the UK Biobank since inception, including enhanced dietary assessment, imaging and assessment of multiple health related outcomes.

Ethical approval for the UK Biobank study was provided by the North West–Haydock Research Ethics Committee (REC reference: 16/NW/0274), and all participants provided electronic signed consent.

### Dietary assessment and calculation of Eatwell Guide scores

Dietary data was collected using a basic touchscreen questionnaire and a more granular assessment instrument, the Oxford WebQ, which is a validated dietary assessment tool for use in large scale observational studies (Greenwood et al., 2019; Liu et al., 2011). The Oxford WebQ is a web-based, self-administered tool which collects data about the consumption of 206 types of foods and 32 types of drinks over a 24-hour period, with participants selecting the number of standard portions for each item consumed. Participants recruited between April 2009 and September 2010 completed the Oxford WebQ as part of their baseline visit and those who provided a valid email address were invited to complete an additional four dietary assessments between February 2011 and June 2012. Dietary reports which were self-reported as atypical by participants were removed from the analysis.

### Eatwell Guide adherence score

We assessed Eatwell Guide adherence using a graded, food-based score modelled on the Panagiotakos Pyramid MedDiet score (Panagiotakos et al., 2007). Building on our previous work (Gregory et al., 2024), this score awarded points for adherence to the Eatwell Guide for the following food (rather than nutrient and food) based components; starchy carbohydrates, wholegrains, beans and pulses, fish, white meat, nuts, eggs, red and processed meat, fruit and vegetables, dairy, discretionary foods and fluid. A list of individual foods which contributed to each food group is available in supplementary material 1. The use of a food-based Eatwell Guide scoring tool aligns more closely with other dietary pattern assessment tools such as the Mediterranean diet (Panagiotakos et al., 2007) and provides clearer public health guidance. In addition, food-based scores facilitate easier quantification of adherence for researchers and participants, and allows for better monitoring of diet (Willett, 2013).

The Eatwell Guide adherence tool awarded 0 to 5 points according to level of compliance with recommendations, with a total possible score of 60 points. The criteria for achieving 0 to 5 points were derived from Eatwell Guide recommendations (PHE, 2016; Scarborough et al., 2016). Five points were awarded if a participant met the Eatwell Guide recommendation for a particular food group. For healthy foods (starchy carbohydrates, wholegrains, beans and pulses, fish, white meat, nuts, eggs, fruit and vegetables, fluid), 0 points were awarded for achieving less than half the recommended intake, whilst for unhealthy foods (red and processed meat, dairy, discretionary foods), 0 points were awarded for consuming 1.5 times the recommended limit. Taking wholegrains as an example, 5 points were awarded for consumption of ≥ 3 servings/d, 4 points for ≥ 2.625-2.999 servings/d, 3 points for ≥ 2.25 – 2.6249 servings/d, 2 points for ≥1.875 – 2.2499 servings/d, 1 point for ≥1.5 – 1.8749 servings/d and 0 points for < 1.5 servings/d. The full scoring methodology is available in supplementary material 1.

### Statistical analysis

All analyses were conducted in SPSS version 28. Baseline characteristics of the analytic sample were summarised as mean ± SD for continuous variables and as number and percentage of participants for categorical variables. Eatwell Guide adherence scores were grouped according to participant age (grouped as ‘younger’ vs. ‘older’ by dichotimizing at the median (57 years)), sex (male, female), BMI (healthy, overweight, obesity), socioeconomic status (Townsend deprivation Index categorised as low [quintile 1], moderate [quintiles 2–4], high [quintile 5] deprivation) (Shannon et al., 2023), education (higher [college/university/other professional qualification], vocational [NVQ/HND/HNC], upper secondary [A-levels], lower secondary [O-levels/GCSEs /CSEs] or none/prefer not to say) and ethnicity (White, Mixed, South Asian, Black, Chinese). Due to the very small number of participants within the underweight category, all participants with a BMI under 25 were categorised within the ‘healthy’ subgroup. Differences in mean total Eatwell Guide adherence scores according to sex, age ethnicity (white vs non-white) were analysed using independent sample t-tests and for BMI, socio-economic status and education using one-way ANOVA and Tukey’s post-hoc test. Due to the small sample size for non-white ethnicities, a grouped white vs non-white comparison was conducted for inferential statistics, however descriptive statistics for all ethnicities are available in relevant tables.

Sociodemographic characteristics by tertiles of adherence to the Eatwell Guide were also analysed to provide information about the distribution of adherence scores. A one-way ANOVA test was used to assess differences in adherence according to age, and associations between adherence tertile and sex, ethnicity, socio-economic status and education using Pearson’s chi^2^ test. We also assessed sociodemographic characteristics by adherence to individual components of the Eatwell Guide via independent sample t-tests, one-way ANOVA and Tukey’s post-hoc test.

Sensitivity analyses were performed to test the robustness of the associations between socio-demographic factors and Eatwell Guide adherence. These involved: 1) only including participants with a minimum of two 24-h diet recalls to provide a more robust measure of habitual diet, 2) excluding participants with 24-h recalls with extreme energy intakes (defined as < 800 or > 4200 kcal/d for males and < 600 or > 3500 kcal/d for females) and 3) assessing whether individual components drive associations by sequentially removing each component from the Eatwell Guide score. Sensitivity analysis were also conducted by further exploring the influence of age on Eatwell Guide adherence, by grouping into older (65 years and over) vs. younger (<65 years) participants using a more common threshold for distinguishing between these groups.

## RESULTS

### Characteristics of UK Biobank participants

A total of 192,825 participants were included in the analysis (Table 1). Participants were mostly white (95.5%) and female (54.9%), and with a mean age of 56 years (range 39 – 72 years). Most participants either had a ‘healthy’ (37.6%) or ‘overweight’ (41.3%) BMI, with a smaller proportion in the ‘obesity’ category (20.9%). Over half of participants (57.8%) were educated to degree or other professional qualification level.

**Table 1.** Socio-demographic characteristics of the analytic sample.

|  | Participants with a total adherence score included in the present analysis (n = 192,825) |
| --- | --- |
| **Age (years)** | 56.2 ± 7.9 |
| **Sex (%)** |  |
| Females | 105880 (54.9) |
| Males | 86945 (45.1) |
| **BMI (kg/m<sup>2</sup>)** |  |
| < 25 | 72413 (37.6) |
| 25 – 29.9 | 79648 (41.3) |
| ≥ 30 | 40233 (20.9) |
| **Ethnicity (%)** |  |
| White | 184080 (95.5) |
| Non-white | 7817 (4.1) |
| Mixed | 895 (0.5) |
| South Asian | 2670 (1.4) |
| Black | 2317 (1.2) |
| Chinese | 548 (0.3) |
| Other | 1387 (0.7) |
| Prefer not to say | 312 (0.2) |
| **Education (%)** |  |
| Higher | 111484 (57.8) |
| Vocational | 22432 (11.6) |
| Upper secondary | 11970 (6.2) |
| Lower secondary | 29094 (15.1) |
| None/prefer not to say | 17769 (9.2) |
| **Socioeconomic status (%)** |  |
| 1 (least deprived) | 41463 (21.5) |
| 2-4 | 119148 (61.8) |
| 5 (most deprived) | 31980 (16.6) |

Data are presented as mean ± SD or n (%)
Data missing: Socioeconomic status 0.1%; BMI 0.2%, Ethnicity 0.2%; Education 0.1%

### Total Eatwell Guide adherence score according to socio-demographic factors

The mean total Eatwell Guide adherence score for all participants was 28.74 ± 8.39 and was higher for older (> 57 years) than younger (≤ 57 years) participants (29.35 ± 8.38 vs. 28.13 ± 8.35; *p* < 0.001) and for female compared with male participants (29.72 ± 8.14 vs. 27.54 ± 8.54; *p* < 0.001) (Table 2). Total adherence scores for BMI sub-groups were all significantly different from each other (all *p* < 0.001), with the highest score achieved by those with a ‘healthy’ weight (29.71 ± 8.44), and the lowest score by those within the ‘obesity’ category (28.79 ± 8.21). Eatwell Guide adherence scores were significantly higher in white participants (28.81 ± 8.40) compared with non-white participants (28.12 ± 8.52) (*p* < 0.001). Eatwell Guide adherence scores differed significantly between education levels (all *p* < 0.001) except for vocational vs. lower secondary education (*p* = 0.18). The highest score were achieved by those with a higher (university degree or other professional qualification) education level (29.61 ± 8.31). Total adherence scores by different tertiles of socio-economic status were all significantly different (all *p* < 0.001), with the highest score achieved by the least deprived participants (29.06 ± 8.28), and the lowest score achieved by the most deprived participants (28.19 ± 8.65).

**Table 2.** Total adherence scores by socio-demographic factors.

|  | **Total score** | **P-value** |
| --- | --- | --- |
| **All participants** | 28.74 ± 8.39 |  |
| **Age** |  | **<0.001** |
| Younger (≤ 57 years) | 28.13 ± 8.35 |  |
| Older (> 57 years) | 29.35 ± 8.38 |  |
| **Sex** |  | **<0.001** |
| Female | 29.72 ± 8.14 |  |
| Male | 27.54 ± 8.53 |  |
| **BMI** |  | **<0.001** |
| < 25 | 29.71 ± 8.44 |  |
| 25 – 29.9 | 28.35 ± 8.33 |  |
| > 30 | 28.79 ± 8.21 |  |
| **Ethnicity** |  | **<0.001** |
| White | 28.81 ± 8.40 |  |
| Non-white | 28.12 ± 8.52 |  |
| Mixed | 28.23 ± 8.85 |  |
| South Asian | 27.92 ± 8.07 |  |
| Black | 26.73 ± 8.71 |  |
| Chinese | 30.91 ± 8.67 |  |
| Other | 29.34 ± 8.52 |  |
| Prefer not to say | 28.03 ± 8.33 |  |
| **Education** |  | **<0.001** |
| Higher | 29.61 ± 8.31 |  |
| Vocational | 27.41 ± 8.32 |  |
| Upper secondary | 28.71 ± 8.49 |  |
| Lower secondary | 27.57 ± 8.29 |  |
| None/prefer not to say | 26.90 ± 8.29 |  |
| **Socio-economic status** |  | **<0.001** |
| 1 (least deprived) | 29.06 ± 8.28 |  |
| 2-4 | 28.78 ± 8.34 |  |
| 5 (most deprived) | 28.19 ± 8.65 |  |

Data are presented as mean ± SD. Data missing: Socioeconomic status 0.1%; BMI 0.2%; Ethnicity 0.2%; Education 0.1%. *P* value for ethnicity is a comparison of white vs non-white participants

### Participant socio-demographic characteristics according to Eatwell Guide adherence score tertiles

The socio-demographic characteristics of participants according to tertile of Eatwell Guide adherence score can be seen in Table 3. The age of participants increased with tertile of adherence to the Eatwell Guide (*p* < 0.001). There was a lower proportion of females in the lowest tertile of adherence compared to males (46.7% vs. 53.3%), and a much higher proportion of females in the highest tertile of adherence compared with males (61.3% vs. 38.7%) (*p* < 0.001). The proportion of those with a ‘healthy’ BMI rose from 32.9% in the lowest tertile to 42.8% in the highest tertile of adherence. There was a gradual decrease in the proportion of participants with a BMI indicating obesity, with increasing tertiles of adherence (lowest: 23.4% vs. highest: 18.1%) (*p* < 0.001). The proportion of white participants increased from 91.1% in the lowest tertile to 92.7% in the highest tertile of adherence, whereas the proportion of non-white participants decreased from 4.3% in the lowest tertile to 3.6% in the highest tertile (p *<* 0.001). The proportion of those with a higher education level (university degree or other professional qualification) rose from 50.8% in the lowest tertile to 64.2% in the highest tertile (*p* < 0.001). The proportion of ‘most deprived’ participants gradually decreased with increasing tertiles of adherence to the Eatwell Guide (lowest: 18.7% vs. highest 15.7%) (*p* < 0.001).

**Table 3.** Sociodemographic characteristics of included participants according to tertiles of Eatwell Guide adherence.

| **Total adherence score tertile** | **Lowest ($\leq 25$)** | **Medium (25.01 – 32)** | **Highest ($>32$)** | **P-value** |
| --- | --- | --- | --- | --- |
| **N** | 61273 | 67324 | 64228 |  |
| **Age (years)** | 55.45 ± 8.15 | 56.23 ± 7.9 | 56.95 ± 7.64 | **<0.001** |
| **Sex** |  |  |  | **<0.001** |
| Female | 28608 (46.7) | 37927 (56.3) | 39345 (61.3) |  |
| Male | 32665 (53.3) | 29397 (43.7) | 24883 (38.7) |  |
| **BMI** |  |  |  | **<0.001** |
| < 25 | 20120 (32.9) | 24882 (37.1) | 27411 (42.8) |  |
| 25 – 29.9 | 26625 (43.6) | 27908 (41.6) | 25088 (39.2) |  |
| > 30 | 14299 (23.4) | 14356 (21.4) | 11578 (18.1) |  |
| **Ethnicity** |  |  |  | **<0.001** |
| White | 58222 (91.1) | 64256 (91.9) | 61602 (92.7) |  |
| Non-white | 2731 (4.3) | 2712 (3.9) | 2374 (3.6) |  |
| Mixed | 316 (0.5) | 294 (0.4) | 285 (0.4) |  |
| South Asian | 893 (1.4) | 1020 (1.5) | 757 (1.14) |  |
| Black | 986 (1.5) | 724 (1.0) | 608 (0.9) |  |
| Chinese | 129 (0.2) | 180 (0.3) | 239 (0.4) |  |
| Other | 408 (0.6) | 494 (0.7) | 484 (0.7) |  |
| Prefer not to say | 205 (0.3) | 241 (0.3) | 170 (0.3) |  |
| **Education** |  |  |  | **<0.001** |
| Higher | 31093 (50.8) | 39173 (58.2) | 41218 (64.2) |  |
| Vocational | 8497 (13.9) | 7732 (11.5) | 6203 (9.7) |  |
| Upper secondary | 3852 (6.3) | 4094 (6.1) | 4024 (6.3) |  |
| Lower secondary | 10664 (17.4) | 10247 (15.2) | 8183 (12.7) |  |
| None/prefer not to say | 7138 (11.7) | 6046 (9) | 4585 (7.1) |  |
| **Socio-economic status** |  |  |  | **<0.001** |
| 1 (least deprived) | 12476 (20.4) | 14744 (21.9) | 14243 (22.2) |  |
| 2-4 | 37671 (61.6) | 41627 (61.9) | 39850 (62.1) |  |
| 5 (most deprived) | 11025 (18) | 10886 (16.2) | 10069 (15.7) |  |

All data presented as n (%) except for age which is presented as mean ± SD. Percentages are rounded to 1 decimal place. Statistical analysis of ethnicity includes white vs non-white participants

### Adherence to individual components of the Eatwell Guide adherence score by socio-demographic factors

Adherence scores to individual components of the Eatwell Guide by socio-demographic factors are presented in supplementary material 2-13 and summarised here. The percentage of participants who achieved full adherence (i.e. 5 points) to individual components of the Eatwell Guide ranged from 15.3% for wholegrains to 72.3% for dairy (supplementary table 14). Adherence scores for starchy carbohydrate, wholegrains, fish, fruit and vegetables, beans and pulses, nuts, egg and discretionary foods was higher in older (> 57 years) than younger (< 57 years) participants, whereas adherence to white meat, dairy and fluid recommendations was higher in younger than older participants (all *p* < 0.001). No significant association was observed between age and red and processed meat scores (*p* = 0.268). Adherence scores for starchy carbohydrate, red and processed meat, fish, fruit and vegetable intake, nuts, discretionary foods and fluid were higher in females than male participants (all *p* < 0.001), whereas adherence scores for wholegrains, dairy and beans and pulses were higher in males than females (all *p* ≤ 0.02). No significant influence of sex was observed on adherence to white meat (*p* = 0.95) or egg recommendations (*p* = 0.08). There was a significant effect of BMI on all components of the Eatwell Guide (all *p* < 0.001) with the exception dairy (*p* = 0.712). Those with a healthy BMI had significantly higher adherence scores for starchy carbohydrate, wholegrain, red and processed meat, fish, fruit and vegetables, beans and pulses and nuts when compared with those in the overweight and/or obesity categories (all *p* < 0.05). Those participants within the obesity BMI category had a higher adherence to white meat, eggs, discretionary food and fluid recommendations than those in the overweight and/or healthy BMI category (*p* < 0.05).

Adherence scores for starchy carbohydrate, wholegrains, fish, fruit and vegetables, beans and pulses, discretionary foods and fluid were higher in white compared with non-white participants (all *p* < 0.001). In contrast, adherence scores were higher in non-white compared with white participants for red and processed meat, dairy and nuts (all *p* < 0.001). No differences were observed for adherence to white meat (*p* = 0.67) or egg recommendations (*p* = 0.74). There was a significant effect of education level on all individual components of the Eatwell Guide (all *p* = 0.001). Notably, those with a higher education level (university degree or other professional qualification) had higher adherence scores for most Eatwell Guide components, including wholegrains, red and processed meat, fish, fruit and vegetables, beans and pulses, nuts and discretionary foods than those of other education levels (all *p* < 0.05). A notable exception was that adherence scores for dairy were lower in those with a higher education level than all other education levels (*p* < 0.05). Other pairwise comparisons can be found in supplementary tables 2-13.

There was a significant effect of socio-economic status on all individual components of the Eatwell Guide (all *p* < 0.001), with the exception of nuts (*p* = 0.39), in which adherence was poor across all groups (∼1 point achieved in each group). Adherence scores for starchy carbohydrate, wholegrains, fish, white meat, fruit and vegetables, beans and pulses, were higher in the least deprived compared with most deprived participants (*p* < 0.05). Adherence scores for red and processed meat, dairy, eggs, discretionary foods and fluid were higher in the most deprived compared with least deprived participants (*p* < 0.05).

### Sensitivity analysis

The associations between Eatwell Guide adherence score and socio-demographic characteristics were robust to a range of sensitivity analyses. Results were similar and remained significant when we repeated the analyses only including those participants who completed a minimum of two dietary reports (with the exception of ethnicity which became non-significant) (supplementary material 15), excluding those with extreme energy intakes (defined as < 800 or > 4200 kcal/d for males and < 600 or > 3500 kcal/d for females) (supplementary material 16), and sequentially removed one component of the Eatwell Guide score (with the exception of ethnicity which became non-significant when starchy carbohydrate was removed) (supplementary material 17). Associations between Eatwell Guide adherence and age remained significant when grouped into older (≥ 65 years) and younger (< 65 years) age categories (*p* < 0.001).

## DISCUSSION

This is the first study to comprehensively evaluate the influence of sociodemographic factors on adherence to the Eatwell Guide (the UK’s healthy eating model). Using a graded, food-based scoring system, we found significant differences in total adherence to the Eatwell Guide between different socio-demographic groups, including different age, sex, BMI, education and socio-economic groups within the UK Biobank cohort. Specifically, older, female participants, and those with a healthy BMI tended to have greater adherence to the Eatwell Guide. In addition, those with a higher education level, and a lower deprivation status had greater adherence to the Eatwell Guide.

The small, albeit significantly higher adherence to the Eatwell Guide in older (> 57 years) vs. younger (≤ 57 years) participants was characterised by greater adherence scores for starchy carbohydrate, wholegrains, fish, fruit and vegetables, beans and pulses, nuts, egg and discretionary foods. These associations remained when we used the more common threshold of ≥65 years to delineate older/younger adults. This is consistent with Carrasco-Marin et al., (2024) who observed that older adults generally had a healthier diet than younger adults. This is also broadly consistent with Malcomson et al. (2023) who also observed higher fruit and vegetable intake and decreased red and processed meat intake in older individuals in the same cohort. Data from the US suggests that diet quality is better in children (2-17 years) and older (65+ years) adults than younger and middle aged (18-65) adults (Hiza et al., 2013). This observation in older adults may be attributed to a more health conscious mindset with increasing age, the adoption of a healthier diet to help management of chronic diseases (Hiza et al., 2013) or time availability to buy, prepare and cook healthy foods (Whitelock and Ensaff, 2018). In contrast, older adults tended to have lower adherence to dairy, which is an important source of macronutrients and micronutrients and is associated with healthy ageing (Lana et al., 2015; Lee et al., 2018). In addition, fluid intake was also lower in older adults, which requires consideration given the prevalence of dehydration in this population (Beck et al., 2021). Given the UK Biobank cohort predominantly comprises middle-aged adults and does not fully reflect the age profile of the UK population (Fry et al., 2017), inferences on diet quality over the life course is limited and requires further evaluation in different UK cohorts.

The higher adherence to the Eatwell Guide observed in female compared with male participants is explained by greater adherence to individual components such as starchy carbohydrate, red and processed meat, fish, fruit and vegetable intake, nuts, discretionary foods and fluid. This is consistent with previous research in this area with females typically demonstrating better overall dietary intake (Carrasco-Marin et al., 2024) and intake of specific food groups (Malcomson et al., 2023). Although all food groups were not reported, Malcomson et al., (2023) did demonstrate greater adherence to fruit and vegetable and red and processed meat recommendations in female compared with male participants in the UK Biobank cohort. Greater intake of fruit and vegetables in females compared with males has been attributed to more favourable attitudes (e.g. perception of health benefits) and greater perceived behaviour control (i.e. ease of facilitating these behaviours) towards fruit and vegetable intake (Emanuel et al., 2012). It may also be plausible that this is attributed to diet culture in women of this age, this weight loss programmes such as Slimming World shown to change behaviours of members to healthier food choices (Pallister et al., 2009). Higher intakes of fruit and vegetables likely infer greater healthy benefits due to their link with reduced with of cardiovascular disease (Tan et al., 2024) and vascular dementia (Griffiths et al., 2024). Interestingly, and consistent with Bennett et al. (2018), we observed that males tended to have greater intake of wholegrains (representative of fibre intake) than females, albeit adherence was generally poor, with just 15.3% of the overall cohort achieving full adherence to wholegrain recommendations. Increasing wholegrain intake by 3 portions per day has been associated with a 25% decreased risk of mortality from cardiovascular disease (Zong et al., 2016), and has also been demonstrated to increase longevity in the UK biobank cohort (Fadnes et al., 2023). As such, public health strategies should be employed to increase intake of wholegrain foods (Boyle et al., 2024).

Higher Eatwell Guide adherence scores were observed in those with a healthy BMI than those with a BMI indicative of obesity. Whilst this finding does suggest that those with a healthy weight typically follow a better diet, some issues of reverse causality may be present (i.e. weight change/status affecting diet rather than diet affecting weight). The current, yet limited evidence base supports adherence to the Eatwell Guide for maintenance of a healthy weight, with Best and Flannery (2023) demonstrating associations with lower waist circumference and lower risk of abdominal obesity in post-menopausal women in the UK women’s cohort study. Despite this, there is a paucity of evidence around the protective effect of adherence to dietary patterns, such as the Eatwell Guide on weight status and future research is required to quantify cross-sectional and prospective associations between Eatwell Guide adherence and weight status in different populations.

Ethnicity was found to be an important factor associated with Eatwell Guide adherence, with white participants demonstrating significantly higher total adherence scores compared to non-white participants. Specifically, white individuals showed better adherence across several food groups, including starchy carbohydrates, wholegrains, fish, fruit and vegetables, beans and pulses, discretionary foods, and fluid intake. These findings align with existing literature indicating that dietary behaviours can vary considerably across ethnic groups, influenced by cultural food preferences, socioeconomic disparities, and varying levels of nutrition knowledge or engagement with dietary guidelines (Ojo et al., 2023a; Ojo et al., 2023b). While lower adherence among non-white groups may reflect broader structural inequalities, it is also important to recognise that many dietary patterns within ethnic minority communities, though not always aligned with the Eatwell Guide, can still be nutritionally beneficial. Public health strategies must be culturally responsive, acknowledging the diversity of dietary practices and barriers across different groups (Bennett et al., 2022). Some culturally adapted versions of the Eatwell Guide have been developed (South Asian (Jay, 2021) and African and Caribbean (Saint Hill, 2023) and could enhance the inclusivity and relevance of future research by coding according to these models (Shannon et al., 2024). Interpretation of these findings are somewhat limited by the high proportion of white participants in the UK Biobank dataset and future work should prioritise more diverse cohorts to further explore these associations. In addition, the significant differences between white and non-white participants became non-significant when only including participants with a minimum of two dietary reports, and when starchy carbohydrate was removed from the total Eatwell Guide score. As such these findings should be interpreted with caution.

Socio-economic status was demonstrated to have an influence on Eatwell Guide adherence, with those least deprived achieving higher scores than the most deprived participants. This is consistent with previous research which has shown similar observations, where individuals in more deprived areas tend to consume fewer wholegrains (Mann et al., 2015), fruit and vegetable (Mackenbach et al., 2015) and fish (Maguire, 2015), and generally have a poorer diet quality (Carrasco-Marin et al., (2024). Recent evidence implicates the food environment, which tends to be more obesogenic in more deprived areas, characterised by a high density of fast-food outlets and limited access to affordable healthy foods (Burgoine et al., 2023; Briazu et al., 2024), both of which have been linked to weight status (Giskes et al., 2011). The ongoing food insecurity crisis in the UK has amplified these disparities, with lower-income households increasingly unable to afford healthy diets (Loopstra et al., 2020). Food price is an important determinant of food choice, with high density energy diets typically associated with lower costs, but also the least healthy option. Healthier foods in the UK, which more closely align with Eatwell Guide recommendations were found to be more expensive than less healthy foods (£0.81/100kcal vs. £0.33/100kcal) (Hoenink et al., 2024). In addition, fruit and vegetables were found to be the most expensive food group in 2023 (Hoenink et al., 2024). These data highlight a potential population-level barrier to adhering to a healthy diet in the UK.

Data from the present study provide numerous practical and research implications for consideration of policy makers, public and researchers alike. A summary of the practical and research recommendations are summarised in table 4. Adherence to the Eatwell Guide is typically poor, with a mean adherence score of only ∼29 out of a maximum 60 points. Adherence to some components are particularly poor, including ∼15% of the population meeting wholegrain intake recommendations, ∼18% meeting fish intake recommendations and ∼20% meeting nut intake recommendations. Given the associated health implications of consuming wholegrains (Barber et al., 2020), fish (Mozaffarian & Rimm, 2006) and nuts (De Souza et al., 2017), public health policy and messaging may benefit from a particular focus on increasing adherence to these food groups. In addition, increased intake of wholegrains and nuts were two of the key factors which led to the largest longevity gains when modelled in the UK Biobank cohorts (Fadnes et al., 2023). Research is required to understand why adherence to the Eatwell Guide is so low in the UK population. Differences in socio-demographic groups were present, although differences in overall Eatwell Guide scores were generally small. This may be a result of the graded (i.e. 0-5 points dependant on adherence), as opposed to binary (i.e. 0 points does not meet recommendations, 1 point meets recommendations) dietary adherence score that was used in this study. Whilst this graded score provides a better reflection of participant diet, the variance in scores is likely to be much lower compared to the use of a cruder, binary score. These small differences may also be due to the homogenous cohort used (likely a result of healthy volunteer bias), and further evaluation using other UK cohorts (e.g., using NDNS data which better reflects the UK population (OHID, 2016)) may be useful. Personalised messaging, which is co-designed with different population subgroups to enhance adherence to the Eatwell Guide is needed to improve dietary intake across the UK.

**Table 4.** Summary of practical/policy recommendations and future research recommendations.

| **Practical/policy recommendations** | **Future research recommendations** |
| --- | --- |
| **Targeted public health messaging** – Develop socio-economically specific public health interventions to address observed disparities in Eatwell Guide adherence | **Conduct prospective analyses on Eatwell Guide adherence and health outcomes** – Investigate the long-term impact of adherence to the Eatwell Guide using prospective designs |
| **Continue to develop inclusive dietary guidelines** – Promote and expand culturally tailored versions of the Eatwell Guide to increase relevance and engagement across diverse ethnic groups | **Utilise more diverse cohort studies** - Conduct studies in more socio-economically diverse (particularly ethnicity and deprivation) UK populations to address the limitations of UK Biobank and better understand specific dietary practices |
| **Develop food specific public health campaigns** – Launch specific campaigns to promote intake of foods such as wholegrains and nuts, given strong links with longevity and disease prevention | **Explore barriers and facilitators to Eatwell Guide adherence** – Conduct quantitative and qualitative research to determine why Eatwell Guide adherence is low across the UK population, with sub-group analysis of socio-economic factors |
| **Engage in community co-design** – Co-create Eatwell Guide adherence interventions with underrepresented or low adherence groups to ensure greater relevance, accessibility and uptake | **Co-create and evaluate interventions/public health campaigns** – Explore the use real world interventions in a range of settings (community, school, workplace, religious centres) aimed at increasing Eatwell Guide adherence |
| **Education based interventions** – Integrate nutrition education into communities, especially those areas with lower educational level given associations with Eatwell Guide adherence |  |

This is the first study to use an advanced, graded score to evaluate adherence to the Eatwell Guide and associations with sociodemographic factors, and has several strengths. These include: the large sample size which maximises statistical power, detailed sociodemographic data which allow us to compare Eatwell Guide scores between various population groups, and our use of several sensitivity analyses to test the assumptions of our statistical approach. Some limitations should be considered. We used dietary data from the Oxford WebQ, a self-administered 24-h recall method, which may be vulnerable to recall and social desirability bias (Naska et al., 2017). Individuals may intentionally misreport intake of particular foods as a result of personal characteristics (e.g. age, sex, BMI) and result in misclassification of dietary intake. For example, those with obesity may be more prone to social desirability bias due to both conscious and unconscious under-reporting (Lissner et al., 2007; Muhlheim et al., 1998). In addition, many participants in the UK Biobank cohort only completed one dietary assessment, and multiple assessments may be required to provide a robust representation of habitual dietary intake (Jackson et al., 2008). Despite these limitations, results from the Oxford WebQ 24-h recall method have been demonstrated to be broadly comparable to interviewer-administered 24-h recalls (Liu et al., 2011) and sensitivity analysis in the present study in which we only included those that completed a minimum of two dietary reports, demonstrated consistent findings. Our findings also build on some other studies (e.g., Carrasco-Marin et al., 2024) which have used the less granular UK Biobank touchscreen questionnaire to measure dietary intake, which lacks information on numerous foods contained within the Eatwell Guide. Further, UK Biobank participants are typically healthier and of higher socio-economic status than the rest of the UK population, which may limit the generalisability of findings. In addition, given participants in the UK Biobank cohort are predominantly white, middle-aged adults, comparison to other ethnic and age groups is somewhat limited.

In conclusion, this large-scale population level cohort study demonstrated that adherence to UK national dietary guidelines differed by socio-demographic factors such as age, sex, BMI, education and socio-economic status. Specifically, those with a higher age, female sex, healthy BMI and higher education and socio-economic status demonstrated greater adherence to the Eatwell Guide. Whilst these data may highlight the requirement for targeted public health messaging to particular sub-groups, poor adherence to the Eatwell Guide across all groups was prevalent, and these findings demonstrate the need for appropriate strategies to increase adherence to the Eatwell Guide.

## Data Availability

All data produced in the present study are available upon reasonable request to the authors

