## Supplementary materials for "Socio-demographic variation in adherence to The Eatwell Guide within the UK Biobank prospective cohort study"

**Supplementary material 1** – List of individual foods which contributed to each food group and the EWG scoring methodology

| Food component | Contributing foods from the Oxford WebQ | Average consumption required. | EWG Score |
|---|---|---|---|
| **Fruit and vegetables** | Stewed fruit (104410), Prune (104420), Dried fruit (104430), Mixed fruit (104440), Apple (104450), Banana (104460), Berry (104470), Cherry (104480), Grapefruit (104490), Grape (104500), Mango (104510), Melon (104520), Orange (104530), Satsuma (104540), Peach nectarine (104550), Pear (104560), Pineapple (104570), Plum (104580), Other fruit (104590), Orange juice (100190), Grapefruit juice (100200), Pure fruit vegetable juice (100210), Fruit smoothie (100220), Mixed veg (104060), Veg pieces (104070), Coleslaw (104080), Side salad (104090), Avocado (104100), Beetroot (104130), Broccoli (104140), Butternut squash (104150), Cabbage kale (104160), Carrot (104170), Cauliflower (104180), Celery (104190), Courgette (104200), Cucumber (104210), Garlic (104220), Leek (104230), Lettuce (104240), Mushroom (104250), Onion (104260), Parsnip (104270), Sweet pepper (104290), Spinach (104300), Sprouts (104310), Sweetcorn (104320), Fresh tomato (104340), Tinned tomato (104350), Turnip swede (104360), Watercress (104370), Other veg (104380), Olive (102490) | From 0 to < 2.5 | 0 |
| | | From 2.5 to < 3.125 | 1 |
| | | From 3.125 to < 3.75 | 2 |
| | | From 3.75 to < 4.375 | 3 |
| | | From 4.375 to < 5 | 4 |
| | | ≥5 | 5 |
| **Starchy Carbohydrates** | White pasta (102710), Wholemeal pasta (102720), White rice (102730), Brown rice (102740), Snackpot (102760), Couscous (102770), Other grain (102780), Sliced bread (100950), Baguette (101020), Bap (101090), Bread roll (101160), Naan bread (101230), Garlic bread (101240), Crispbread (101250), Oatcake (101260), Other bread (101270), Porridge (100770), Muesli (100800), Oat crunch (100810), Plain cereal (100830), Bran cereal (100840), Wholewheat cereal (100850), Other cereal (100860), Fried potatoes (104020), Boiled baked potatoes (104030), Mashed potato (104050), Sweet potato (104330) | **Men**<br>< 2.5<br>**Women**<br>< 2 | 0 |
| | | **Men**<br>From 2.5 to < 3.125<br>**Women**<br>From 2 to < 2.5 | 1 |

| | | | |
|---|---|---|---|
| | | **Men**<br>From 3.125 to < 3.75<br>**Women**<br>From 2.5 to < 3 | 2 |
| | | **Men**<br>From 3.75 to < 4.375<br>**Women**<br>From 3 to < 3.5 | 3 |
| | | **Men**<br>From 4.375 to < 5<br>**Women**<br>From 3.5 to < 4 | 4 |
| | | **Men**<br>≥ 5<br>**Women**<br>≥ 4 | 5 |
| **Wholegrains** | Wholemeal pasta (102720), Brown rice (102740), Sliced bread (100950), Type of bread (20091), Baguette (101020), Type of baguette (20092), Bap (101090), Type of bap (20093), Bread roll (101160), Type of bread roll (20094), Oatcake (101260), Porridge (100770), Muesli (100800), Bran cereal (100840), Wholewheat cereal (100850), Other grain (102780) | From 0 to < 1.5 | 0 |
| | | From 1.5 to < 1.875 | 1 |
| | | From 1.875 to < 2.25 | 2 |
| | | From 2.25 to < 2.625 | 3 |

| | | | |
|---|---|---|---|
| | | From 2.625 to < 3 | 4 |
| | | ≥ 3 | 5 |
| **Beans/pulses** | Pea (104280), Green beans (104120), Broad beans (104110), Baked beans (104000), Pulses (104010), Tofu (103270) | From 0 to < 0.21 | 0 |
| | | From 0.21 to < 0.27 | 1 |
| | | From 0.27 to < 0.32 | 2 |
| | | From 0.32 to < 0.38 | 3 |
| | | From 0.38 to < 0.43 | 4 |
| | | ≥ 0.43 | 5 |
| **Fish**[a] | Tinned tuna (103150), Oily fish (103160), Breaded fish (103170), Battered fish (103180), White fish (103190), Prawns (103200), Lobster crab (103210), Shellfish (103220), Other fish (103230) | **Fish**<br>From 0 to < 0.14<br>**Oily fish**<br>From 0 to < 0.07 | 0 |
| | | **Fish**<br>From 0.14 to < 0.18<br>**Oily fish**<br>From 0.07 to < 0.09 | 1 |
| | | **Fish**<br>From 0.18 to < 0.21<br>**Oily fish**<br>From 0.09 to < 0.11 | 2 |

| | | | |
|---|---|---|---|
| | | **Fish**<br>From 0.21 to < 0.25<br>**Oily fish**<br>From 0.11 to < 0.13 | 3 |
| | | **Fish**<br>From 0.25 to < 0.29<br>**Oily fish**<br>From 0.13 to < 0.14 | 4 |
| | | **Fish**<br>≥ 0.29<br>**Oily fish**<br>≥ 0.14 | 5 |
| **Poultry** | Poultry (103060), Breaded poultry (103050) | From 0 to < 0.07 | 0 |
| | | From 0.07 to < 0.09 | 1 |
| | | From 0.09 to < 0.11 | 2 |
| | | From 0.11 to < 0.13 | 3 |
| | | From 0.13 to < 0.14 | 4 |
| | | ≥ 0.14 | 5 |
| **Nuts** | Unsalted nuts (102440), salted nuts (102430), unsalted peanuts (102420), salted peanuts (102410) | From 0 to < 0.07 | 0 |
| | | From 0.07 to < 0.09 | 1 |

| | | | |
|---|---|---|---|
| | | From 0.09 to < 0.11 | 2 |
| | | From 0.11 to < 0.13 | 3 |
| | | From 0.13 to < 0.14 | 4 |
| | | ≥ 0.14 | 5 |
| **Eggs** | Whole egg (102940), Omelette (102950), Egg sandwiches (102960), Scotch egg (102970), Other egg (102980) | From 0 to < 0.07 | 0 |
| | | From 0.07 to < 0.09 | 1 |
| | | From 0.09 to < 0.11 | 2 |
| | | From 0.11 to < 0.13 | 3 |
| | | From 0.13 to < 0.14 | 4 |
| | | ≥ 0.14 | 5 |
| **Red and processed meat** | Bacon (103070), Ham (103080), Liver (103090), Sausage (103010), Beef (103020), Pork (103030), Lamb (103040) | ≥ 1.5 | 0 |
| | | From 1.375 to < 1.5 | 1 |
| | | From 1.25 to < 1.375 | 2 |
| | | From 1.12 To < 1.25 | 3 |
| | | From 1 to < 1.12 | 4 |
| | | < 1 | 5 |
| **Dairy** | Yogurt (102090), Low fat hard cheese (102810), Hard cheese (102820), Low fat cheese spread (102850), Cheese spread (102860), Soft cheese (102830), Goat cheese (102900), Blue cheese (102840), Feta (102880), Mozzarella (102890), Other cheese (102910), Cottage cheese (102870), Milk (100520), Flavoured milk (100530), Added milk instant coffee (100260), Added milk filtered coffee (100280), Added milk espresso (100320), Added milk other coffee (100350), Added milk standard tea (100460), Added milk rooibos | ≥ 3 | 0 |
| | | From 2.75to < 3 | 1 |
| | | From 2.5 to < 2.75 | 2 |
| | | From 2.25 to < 2.5 | 3 |

| | | | |
|---|---|---|---|
| | tea (100480), Instant coffee (100250), Filtered coffee (100270), Espresso (100310), Other coffee (100330), Standard tea (100400), Rooibos tea (100410), Dairy smoothie (100230), Latte (100300), Cappuccino (100290), Type of milk used (100920), Added milk cereal (100890), Porridge (100770), Muesli (100800), Bran cereal (100840), Wholewheat cereal (100850), Oat crunch (100810), Plain cereal (100830), Sweetened cereal (100820), Other cereal (100860) | From 2 to < 2.25 | 4 |
| | | < 2 | 5 |
| **Discretionary foods** | Chocolate biscuit (102350), Chocolate covered biscuit (102340), Chocolate bar (102260), Chocolate sweets (102310), Chocolate raisins (102300), Dark chocolate (102290), Milk chocolate (102280), White chocolate (102270), Sweet biscuits (102360), Cakes (102190), Cheesecake (102220), Doughnut (102200), Fruitcake (102180), Danish pastry (102060), Sponge pudding (102210), Milk based pudding (102140), Other milk pudding (102150), Other desert (102230), Soya desert (102170), Sweets (102330), Diet sweets (102320), Other sweets (102380), Ice cream (102120), Fizzy drinks (100170), Squash (100180), Sugar added to tea (100490), Sugar added to coffee (100370), Sugar added to cereal (100900), Hot chocolate (100550), Pancake (102010), Scotch pancake (102020), Croissant (102050), Scone (102070), Crisp (102460), Cereal bar (102370) | ≥ 1.5 | 0 |
| | | From 1.375 to < 1.5 | 1 |
| | | From 1.25 to < 1.375 | 2 |
| | | From 1.12 to < 1.25 | 3 |
| | | From 1 to < 1.12 | 4 |
| | | < 1 | 5 |
| **Fluid** | Drinking water (100150), Instant coffee (100250), Filtered coffee (100270), Espresso (100310), Cappuccino (100290), Latte (100300), Other coffee type (100330), Decaffeinated coffee (100360), Standard tea (100400), Rooibos tea (100410), Green tea (100420), Herbal tea (100430), Other tea (100440), Decaffeinated tea (100470), Low calorie drink (100160), Squash (100180), Orange juice (100190), Grapefruit juice (100200), Pure fruit vegetable juice (100210), Fruit smoothie (100220), Dairy smoothie (100230), Type of milk used (100920), Milk (100520), Flavoured milk (100530) | From 0 to < 3 | 0 |
| | | From 3 to < 3.75 | 1 |
| | | From 3.75 to < 4.5 | 2 |
| | | From 4.5 to < 5.25 | 3 |
| | | From 5.25 to < 6 | 4 |
| | | ≥ 6 | 5 |

a; The final EWG score for fish was created by averaging the EWG score for oily fish and the EWG score for fish overall.

**Supplementary material 2** – Starchy carbohydrate intake by socio-demographic factors

| | Total score | P-value |
|---|---|---|
| **All participants** | 3.15 ± 1.83 | |
| **Age** | | **< 0.001** |
| Younger (≤ 57 years) | 3.07 ± 1.87 | |
| Older (> 57 years) | 3.24 ± 1.80 | |
| **Sex** | | **< 0.001** |
| Female | 3.29 ± 1.79 | |
| Male | 2.98 ± 1.87 | |
| **BMI** | | **<0.001** |
| < 25 | 3.28 ± 1.79 | |
| 25 – 29.9 | 3.07 ± 1.85 | |
| > 30 | 3.10 ± 1.87 | |
| **Ethnicity** | | **<0.001** |
| White | 3.18 ± 1.82 | |
| Non-white | 2.62 ± 1.98 | |
| Mixed | 2.98 ± 1.89 | |
| South Asian | 2.50 ± 1.98 | |
| Black | 2.51 ± 2.02 | |
| Chinese | 2.64 ± 1.93 | |
| Other | 2.78 ± 1.93 | |
| Prefer not to say | 2.97 ± 1.85 | |
| **Education** | | **< 0.001** |
| Higher | 3.15 ± 1.83 | |
| Vocational | 3.11 ± 1.85 | |
| Upper secondary | 3.11 ± 1.85 | |
| Lower secondary | 3.16 ± 1.83 | |
| None/prefer not to say | 3.22 ± 1.85 | |
| **Socio-economic status** | | **<0.001** |
| 1 (least deprived) | 3.19 ± 1.81 | |
| 2-4 | 3.17 ± 1.83 | |
| 5 (most deprived) | 3.04 ± 1.89 | |

*P* value for ethnicity in relation to white vs non white participants

**Supplementary material 3** – Wholegrain intake by socio-demographic factors

| | Total score | P-value |
|---|---|---|
| **All participants** | 1.33 ± 1.87 | |
| **Age** | | **< 0.001** |
| Younger (≤ 57 years) | 1.22 ± 1.83 | |
| Older (> 57 years) | 1.44 ± 1.92 | |
| **Sex** | | **< 0.001** |
| Female | 1.14 ± 1.75 | |
| Male | 1.57 ± 2.00 | |
| **BMI** | | **<0.001** |
| < 25 | 1.48 ± 1.93 | |
| 25 – 29.9 | 1.30 ± 1.86 | |
| > 30 | 1.14 ± 1.79 | |
| **Ethnicity** | | **<0.001** |
| White | 1.35 ± 1.88 | |
| Non-white | 1.08 ± 1.78 | |
| Mixed | 1.24 ± 1.88 | |
| South Asian | 1.08 ± 1.77 | |
| Black | 1.06 ± 1.79 | |
| Chinese | 0.87 ± 1.64 | |
| Other | 1.07 ± 1.76 | |
| Prefer not to say | 1.31 ± 1.85 | |
| **Education** | | **<0.001** |
| Higher | 1.42 ± 1.91 | |
| Vocational | 1.25 ± 1.85 | |
| Upper secondary | 1.28 ± 1.84 | |
| Lower secondary | 1.14 ± 1.78 | |
| None/prefer not to say | 1.22 ± 1.84 | |
| **Socio-economic status** | | **<0.001** |
| 1 (least deprived) | 1.37 ± 1.88 | |
| 2-4 | 1.34 ± 1.87 | |
| 5 (most deprived) | 1.28 ± 1.88 | |

*P* value for ethnicity in relation to white vs non white participants

**Supplementary material 4** – Red and processed meat intake by socio-demographic factors

| | Total score | P-value |
|---|---|---|
| **All participants** | 3.65 ± 1.98 | |
| **Age** | | 0.268 |
| Younger (≤ 57 years) | 3.66 ± 1.99 | |
| Older (> 57 years) | 3.65 ± 1.97 | |
| **Sex** | | **<0.001** |
| Female | 3.87 ± 1.85 | |
| Male | 3.39 ± 2.11 | |
| **BMI** | | **<0.001** |
| < 25 | 3.90 ± 1.84 | |
| 25 – 29.9 | 3.59 ± 2.01 | |
| > 30 | 3.35 ± 2.12 | |
| **Ethnicity** | | **<0.001** |
| White | 3.64 ± 1.99 | |
| Non-white | 4.13 ± 1.71 | |
| Mixed | 3.82 ± 1.90 | |
| South Asian | 4.45 ± 1.40 | |
| Black | 3.98 ± 1.83 | |
| Chinese | 3.73 ± 1.94 | |
| Other | 4.13 ± 1.69 | |
| Prefer not to say | 3.77 ± 1.95 | |
| **Education** | | **< 0.001** |
| Higher | 3.72 ± 1.95 | |
| Vocational | 3.50 ± 2.06 | |
| Upper secondary | 3.65 ± 1.99 | |
| Lower secondary | 3.59 ± 2.01 | |
| None/prefer not to say | 3.58 ± 2.00 | |
| **Socio-economic status** | | **<0.001** |
| 1 (least deprived) | 3.62 ± 1.99 | |
| 2-4 | 3.65 ± 1.98 | |
| 5 (most deprived) | 3.72 ± 1.98 | |

*P* value for ethnicity in relation to white vs non white participants

**Supplementary material 5** – Fish intake by socio-demographic factors

| | **Total score** | **P-value** |
|---|---|---|
| **All participants** | 1.53 ± 1.95 | |
| **Age** | | **< 0.001** |
| Younger (≤ 57 years) | 1.41 ± 1.90 | |
| Older (> 57 years) | 1.64 ± 2.00 | |
| **Sex** | | **< 0.001** |
| Female | 1.60 ± 1.98 | |
| Male | 1.44 ± 1.92 | |
| **BMI** | | **< 0.001** |
| < 25 | 1.67 ± 2.01 | |
| 25 – 29.9 | 1.49 ± 1.94 | |
| > 30 | 1.34 ± 1.85 | |
| **Ethnicity** | | **<0.001** |
| White | 1.53 ± 1.96 | |
| Non-white | 1.38 ± 1.92 | |
| Mixed | 1.42 ± 1.90 | |
| South Asian | 0.99 ± 1.68 | |
| Black | 1.54 ± 2.00 | |
| Chinese | 1.99 ± 2.05 | |
| Other | 1.60 ± 2.02 | |
| Prefer not to say | 1.43 ± 1.93 | |
| **Education** | | **< 0.001** |
| Higher | 1.66 ± 2.00 | |
| Vocational | 1.32 ± 1.85 | |
| Upper secondary | 1.56 ± 1.96 | |
| Lower secondary | 1.34 ± 1.86 | |
| None/prefer not to say | 1.21 ± 1.80 | |
| **Socio-economic status** | | **< 0.001** |
| 1 (least deprived) | 1.60 ± 1.98 | |
| 2-4 | 1.52 ± 1.95 | |
| 5 (most deprived) | 1.45 ± 1.92 | |

*P* value for ethnicity in relation to white vs non white participants

**Supplementary material 6** – White meat intake by socio-demographic factors

| | **Total score** | **P-value** |
|---|---|---|
| **All participants** | 2.00 ± 2.44 | |
| **Age** | | **< 0.001** |
| Younger (≤ 57 years) | 2.07 ± 2.46 | |
| Older (> 57 years) | 1.94 ± 2.43 | |
| **Sex** | | 0.95 |
| Female | 2.01 ± 2.44 | |
| Male | 2.01 ± 2.44 | |
| **BMI** | | **< 0.001** |
| < 25 | 1.95 ± 2.43 | |
| 25 – 29.9 | 2.03 ± 2.45 | |
| > 30 | 2.06 ± 2.44 | |
| **Ethnicity** | | **0.64** |
| White | 2.01 ± 2.44 | |
| Non-white | 2.02 ± 2.45 | |
| Mixed | 1.95 ± 2.44 | |
| South Asian | 1.69 ± 2.36 | |
| Black | 2.33 ± 2.49 | |
| Chinese | 2/33 ± 2.49 | |
| Other | 2.05 ± 2.46 | |
| Prefer not to say | 1.82 ± 2.40 | |
| **Education** | | **< 0.001** |
| Higher | 2.01 ± 2.44 | |
| Vocational | 2.09 ± 2.46 | |
| Upper secondary | 2.09 ± 2.46 | |
| Lower secondary | 2.03 ± 2.45 | |
| None/prefer not to say | 1.80 ± 2.40 | |
| **Socio-economic status** | | **< 0.001** |
| 1 (least deprived) | 2.08 ± 2.46 | |
| 2-4 | 2.01 ± 2.44 | |
| 5 (most deprived) | 1.89 ± 2.42 | |

*P* value for ethnicity in relation to white vs non white participants

**Supplementary material 7** – Fruit and vegetable intake by socio-demographic factors

| | Total score | P-value |
|---|---|---|
| **All participants** | 3.25 ± 2.00 | |
| **Age** | | **< 0.001** |
| Younger (≤ 57 years) | 3.05 ± 2.06 | |
| Older (> 57 years) | 3.44 ± 1.93 | |
| **Sex** | | **< 0.001** |
| Female | 3.52 ± 1.91 | |
| Male | 2.91 ± 2.07 | |
| **BMI** | | **< 0.001** |
| < 25 | 3.43 ± 1.93 | |
| 25 – 29.9 | 3.18 ± 2.02 | |
| > 30 | 3.05 ± 2.08 | |
| **Ethnicity** | | **<0.001** |
| White | 3.26 ± 2.00 | |
| Non-white | 3.01 ± 2.10 | |
| Mixed | 3.13 ± 2.07 | |
| South Asian | 2.91 ± 2.11 | |
| Black | 2.81 ± 2.16 | |
| Chinese | 3.28 ± 1.96 | |
| Other | 3.36 ± 2.02 | |
| Prefer not to say | 3.12 ± 2.04 | |
| **Education** | | **< 0.001** |
| Higher | 3.44 ± 1.92 | |
| Vocational | 2.94 ± 2.09 | |
| Upper secondary | 3.19 ± 2.02 | |
| Lower secondary | 2.99 ± 2.08 | |
| None/prefer not to say | 2.86 ± 2.12 | |
| **Socio-economic status** | | **< 0.001** |
| 1 (least deprived) | 3.33 ± 1.96 | |
| 2-4 | 3.27 ± 2.00 | |
| 5 (most deprived) | 3.07 ± 2.08 | |

*P* value for ethnicity in relation to white vs non white participants

**Supplementary material 8** – Dairy intake by socio-demographic factors

| | Total score | P-value |
|---|---|---|
| **All participants** | 4.21 ± 1.52 | |
| **Age** | | **< 0.001** |
| Younger (≤ 57 years) | 4.24 ± 1.50 | |
| Older (> 57 years) | 4.18 ± 1.53 | |
| **Sex** | | **< 0.001** |
| Female | 4.20 ± 1.52 | |
| Male | 4.22 ± 1.51 | |
| **BMI** | | 0.712 |
| < 25 | 4.21 ± 1.51 | |
| 25 – 29.9 | 4.21 ± 1.51 | |
| > 30 | 4.20 ± 1.54 | |
| **Ethnicity** | | **<0.001** |
| White | 4.20 ± 1.52 | |
| Non-white | 4.49 ± 1.31 | |
| Mixed | 4.31 ± 1.48 | |
| South Asian | 4.49 ± 1.30 | |
| Black | 4.60 ± 1.19 | |
| Chinese | 4.75 ± 0.93 | |
| Other | 4.34 ± 1.47 | |
| Prefer not to say | 4.22 ± 1.54 | |
| **Education** | | **< 0.001** |
| Higher | 4.18 ± 1.54 | |
| Vocational | 4.24 ± 1.50 | |
| Upper secondary | 4.24 ± 1.50 | |
| Lower secondary | 4.25 ± 1.48 | |
| None/prefer not to say | 4.26 ± 1.48 | |
| **Socio-economic status** | | **< 0.001** |
| 1 (least deprived) | 4.18 ± 1.53 | |
| 2-4 | 4.21 ± 1.51 | |
| 5 (most deprived) | 4.24 ± 1.52 | |

*P* value for ethnicity in relation to white vs non white participants

**Supplementary material 9** – Beans and pulses intake by socio-demographic factors

| | Total score | P-value |
|---|---|---|
| **All participants** | 2.02 ± 2.33 | |
| **Age** | | **< 0.001** |
| Younger (≤ 57 years) | 1.94 ± 2.32 | |
| Older (> 57 years) | 2.11 ± 2.35 | |
| **Sex** | | **0.02** |
| Female | 2.01 ± 2.33 | |
| Male | 2.04 ± 2.34 | |
| **BMI** | | **< 0.001** |
| < 25 | 2.07 ± 2.33 | |
| 25 – 29.9 | 2.02 ± 2.34 | |
| > 30 | 1.97 ± 2.33 | |
| **Ethnicity** | | **< 0.001** |
| White | 2.03 ± 2.34 | |
| Non-white | 1.88 ± 2.32 | |
| Mixed | 1.95 ± 2.32 | |
| South Asian | 2.26 ± 2.39 | |
| Black | 1.46 ± 2.18 | |
| Chinese | 1.83 ± 2.28 | |
| Other | 1.80 ± 2.29 | |
| Prefer not to say | 2.02 ± 2.34 | |
| **Education** | | **< 0.001** |
| Higher | 2.08 ± 2.34 | |
| Vocational | 1.96 ± 2.33 | |
| Upper secondary | 1.98 ± 2.33 | |
| Lower secondary | 1.96 ± 2.33 | |
| None/prefer not to say | 1.91 ± 2.33 | |
| **Socio-economic status** | | **< 0.001** |
| 1 (least deprived) | 2.06 ± 2.34 | |
| 2-4 | 2.04 ± 2.34 | |
| 5 (most deprived) | 1.94 ± 2.33 | |

*P* value for ethnicity in relation to white vs non white participants

**Supplementary material 10** – Nuts intake by socio-demographic factors

| | Total score | P-value |
|---|---|---|
| **All participants** | 1.00 ± 1.99 | |
| **Age** | | **< 0.001** |
| Younger (≤ 57 years) | 0.95 ± 1.95 | |
| Older (> 57 years) | 1.05 ± 2.02 | |
| **Sex** | | **< 0.001** |
| Female | 1.04 ± 2.01 | |
| Male | 0.96 ± 1.96 | |
| **BMI** | | **< 0.001** |
| < 25 | 1.17 ± 0.97 | |
| 25 – 29.9 | 0.97 ± 1.96 | |
| > 30 | 0.76 ± 1.78 | |
| **Ethnicity** | | **<0.001** |
| White | 0.99 ± 1.98 | |
| Non-white | 1.28 ± 2.17 | |
| Mixed | 1.19 ± 2.12 | |
| South Asian | 1.43 ± 2.25 | |
| Black | 1.03 ± 2.02 | |
| Chinese | 1.77 ± 2.37 | |
| Other | 1.28 ± 2.16 | |
| Prefer not to say | 0.98 ± 1.97 | |
| **Education** | | **< 0.001** |
| Higher | 1.11 ± 2.06 | |
| Vocational | 0.85 ± 1.87 | |
| Upper secondary | 1.02 ± 2.00 | |
| Lower secondary | 0.87 ± 1.88 | |
| None/prefer not to say | 0.73 ± 1.75 | |
| **Socio-economic status** | | **0.387** |
| 1 (least deprived) | 1.01 ± 1.99 | |
| 2-4 | 1.00 ± 1.93 | |
| 5 (most deprived) | 1.01 ± 2.00 | |

*P* value for ethnicity in relation to white vs non white participants

**Supplementary material 11** – Egg intake by socio-demographic factors

| | Total score | P-value |
|---|---|---|
| **All participants** | 1.64 ± 2.35 | |
| **Age** | | **< 0.001** |
| Younger (≤ 57 years) | 1.54 ± 2.31 | |
| Older (> 57 years) | 1.74 ± 2.38 | |
| **Sex** | | 0.08 |
| Female | 1.64 ± 2.35 | |
| Male | 1.63 ± 2.34 | |
| **BMI** | | **< 0.001** |
| < 25 | 1.58 ± 2.32 | |
| 25 – 29.9 | 1.64 ± 2.34 | |
| > 30 | 1.74 ± 2.38 | |
| **Ethnicity** | | 0.74 |
| White | 1.64 ± 2.35 | |
| Non-white | 1.65 ± 2.35 | |
| Mixed | 1.57 ± 2.32 | |
| South Asian | 1.49 ± 2.29 | |
| Black | 1.54 ± 2.31 | |
| Chinese | 2.41 ± 2.50 | |
| Other | 1.87 ± 2.42 | |
| Prefer not to say | 1.73 ± 2.38 | |
| **Education** | | **< 0.001** |
| Higher | 1.65 ± 2.35 | |
| Vocational | 1.60 ± 2.33 | |
| Upper secondary | 1.66 ± 2.35 | |
| Lower secondary | 1.60 ± 2.33 | |
| None/prefer not to say | 1.66 ± 2.36 | |
| **Socio-economic status** | | **< 0.001** |
| 1 (least deprived) | 1.60 ± 2.33 | |
| 2-4 | 1.63 ± 2.34 | |
| 5 (most deprived) | 1.72 ± 2.37 | |

*P* value for ethnicity in relation to white vs non white participants

**Supplementary material 12** – Discretionary foods intake by socio-demographic factors

| | Total score | P-value |
|---|---|---|
| **All participants** | 1.40 ± 2.18 | |
| **Age** | | **< 0.001** |
| Younger (≤ 57 years) | 1.37 ± 2.17 | |
| Older (> 57 years) | 1.43 ± 2.20 | |
| **Sex** | | **< 0.001** |
| Female | 1.58 ± 2.25 | |
| Male | 1.18 ± 2.08 | |
| **BMI** | | **< 0.001** |
| < 25 | 1.41 ± 2.18 | |
| 25 – 29.9 | 1.37 ± 2.17 | |
| > 30 | 1.44 ± 2.22 | |
| **Ethnicity** | | **<0.001** |
| White | 1.40 ± 2.18 | |
| Non-white | 1.33 ± 2.16 | |
| Mixed | 1.26 ± 2.12 | |
| South Asian | 1.28 ± 2.14 | |
| Black | 1.14 ± 2.06 | |
| Chinese | 1.79 ± 2.33 | |
| Other | 1.60 ± 2.28 | |
| Prefer not to say | 1.34 ± 2.18 | |
| **Education** | | **< 0.001** |
| Higher | 1.50 ± 2.22 | |
| Vocational | 1.20 ± 2.08 | |
| Upper secondary | 1.40 ± 2.18 | |
| Lower secondary | 1.27 ± 2.12 | |
| None/prefer not to say | 1.27 ± 2.13 | |
| **Socio-economic status** | | **< 0.001** |
| 1 (least deprived) | 1.38 ± 2.17 | |
| 2-4 | 1.39 ± 2.18 | |
| 5 (most deprived) | 1.45 ± 2.21 | |

*P* value for ethnicity in relation to white vs non white participants

**Supplementary material 13** – Fluid intake by socio-demographic factors

| | Total score | P-value |
|---|---|---|
| **All participants** | 3.53 ± 1.76 | |
| **Age** | | **< 0.001** |
| Younger (≤ 57 years) | 3.59 ± 1.75 | |
| Older (> 57 years) | 3.47 ± 1.77 | |
| **Sex** | | **< 0.001** |
| Female | 3.80 ± 1.61 | |
| Male | 3.20 ± 1.88 | |
| **BMI** | | **< 0.001** |
| < 25 | 1.58 ± 2.32 | |
| 25 – 29.9 | 1.64 ± 2.34 | |
| > 30 | 1.74 ± 2.38 | |
| **Ethnicity** | | **< 0.001** |
| White | 3.55 ± 1.76 | |
| Non-white | 3.18 ± 1.90 | |
| Mixed | 3.38 ± 1.86 | |
| South Asian | 3.36 ± 1.82 | |
| Black | 2.68 ± 1.99 | |
| Chinese | 3.50 ± 1.78 | |
| Other | 3.39 ± 1.80 | |
| Prefer not to say | 3.32 ± 1.88 | |
| **Education** | | **< 0.001** |
| Higher | 1.65 ± 2.35 | |
| Vocational | 1.60 ± 2.33 | |
| Upper secondary | 1.66 ± 2.35 | |
| Lower secondary | 1.60 ± 2.33 | |
| None/prefer not to say | 1.66 ± 2.36 | |
| **Socio-economic status** | | **< 0.001** |
| 1 (least deprived) | 1.60 ± 2.33 | |
| 2-4 | 1.63 ± 2.34 | |
| 5 (most deprived) | 1.72 ± 2.37 | |

*P* value for ethnicity in relation to white vs non white participants

**Supplementary table 14**. Proportion of participants achieving adherence to individual components (i.e. 5 points) of the Eatwell guide.

| | **Participants achieving full adherence (N (%))** |
|---|---|
| **Starchy carbohydrates** | 75574 (39.2) |
| **Wholegrains** | 29470 (15.3) |
| **Red and processed meat** | 108600 (56.3) |
| **Fish** | 34432 (17.9) |
| **White meat** | 76756 (39.8) |
| **Fruit and vegetables** | 93877 (48.7) |
| **Dairy** | 139460 (72.3) |
| **Beans and pulses** | 68868 (35.7) |
| **Nuts** | 37587 (19.5) |
| **Eggs** | 63065 (32.7) |
| **Discretionary foods** | 49847 (25.9) |
| **Fluid** | 91115 (47.3) |

**Supplementary table 15**. Sensitivity analysis demonstrating adherence to the Eatwell guide by socio-demographic characteristics, only including those participants who completed a minimum of two dietary reports

| | **Total score** | **P-value** |
|---|---|---|
| **All participants** | | |
| **Age** | | **<0.001** |
| Younger (≤ 57 years) | 29.61 ± 8.24 | |
| Older (> 57 years) | 31.01 ± 8.26 | |
| **Sex** | | **<0.001** |
| Female | 31.25 ± 8.03 | |
| Male | 29.10 ± 8.43 | |
| **BMI** | | **<0.001** |
| < 25 | 31.19 ± 8.29 | |
| 25 – 29.9 | 29.90 ± 8.25 | |
| > 30 | 29.34 ± 8.14 | |
| **Ethnicity** | | 0.93 |
| White | 30.3 ± 8.3 | |
| Non-white | 30.3 ± 8.4 | |
| Mixed | 29.8 ± 8.6 | |
| South Asian | 30.2 ± 7.8 | |
| Black | 29.0 ± 8.8 | |
| Chinese | 32.4 ± 8.7 | |
| Other | 31.5 ± 8.4 | |
| Prefer not to say | 29.6 ± 8.2 | |
| **Education** | | **<0.001** |
| Higher | 30.88 ± 8.22 | |
| Vocational | 29.04 ± 8.27 | |
| Upper secondary | 30.18 ± 8.39 | |
| Lower secondary | 29.22 ± 8.23 | |
| None/prefer not to say | 29.27 ± 8.25 | |
| **Socio-economic status** | | **<0.001** |
| 1 (least deprived) | 30.52 ± 8.17 | |
| 2-4 | 30.38 ± 8.25 | |
| 5 (most deprived) | 29.98 ± 8.53 | |

*P* value for ethnicity in relation to white vs non white participants

**Supplementary table 16**. Sensitivity analysis demonstrating adherence to the Eatwell guide by socio-demographic characteristics, excluding dietary reports with extreme energy intakes

| | **Total score** | **P-value** |
|---|---|---|
| **All participants** | | |
| **Age** | | **<0.001** |
| Younger (≤ 57 years) | 28.09 ± 8.34 | |
| Older (> 57 years) | 29.33 ± 8.37 | |
| **Sex** | | **<0.001** |
| Female | 29.68 ± 8.13 | |
| Male | 27.51 ± 8.51 | |
| **BMI** | | **<0.001** |
| < 25 | 29.67 ± 8.44 | |
| 25 – 29.9 | 28.32 ± 8.32 | |
| > 30 | 27.75 ± 8.19 | |
| **Ethnicity** | | **<0.001** |
| White | 28.76 ± 8.40 | |
| Non-white | 28.05 ± 8.51 | |
| Mixed | 28.18 ± 8.83 | |
| South Asian | 27.91 ± 8.05 | |
| Black | 26.72 ± 8.66 | |
| Chinese | 30.88 ± 8.66 | |
| Other | 29.28 ± 8.48 | |
| Prefer not to say | 28.01 ± 8.31 | |
| **Education** | | **<0.001** |
| Higher | 29.57 ± 8.30 | |
| Vocational | 27.37 ± 8.31 | |
| Upper secondary | 28.67 ± 8.48 | |
| Lower secondary | 27.55 ± 8.28 | |
| None/prefer not to say | 26.87 ± 8.26 | |
| **Socio-economic status** | | **<0.001** |
| 1 (least deprived) | 29.02 ± 8.27 | |
| 2-4 | 28.74 ± 8.33 | |
| 5 (most deprived) | 28.16 ± 8.63 | |

**Supplementary table 17.** Sensitivity analysis of EWG adherence score by socio-demographic factors when sequentially removing one component of EWG

| | Full score | Minus starchy carbohy-drate | Minus wholegrains | Minus red and processed meat | Minus fish | Minus white meat | Minus fruit and veg | Minus dairy | Minus beans and pulses | Minus nuts | Minus egg | Minus discreti-onary food | Minus fluid |
|---|---|---|---|---|---|---|---|---|---|---|---|---|---|
| **Age** | **< 0.001** | **< 0.001** | **< 0.001** | **< 0.001** | **< 0.001** | **< 0.001** | **< 0.001** | **< 0.001** | **< 0.001** | **< 0.001** | **< 0.001** | **< 0.001** | **< 0.001** |
| **Sex** | **< 0.001** | **< 0.001** | **< 0.001** | **< 0.001** | **< 0.001** | **< 0.001** | **< 0.001** | **< 0.001** | **< 0.001** | **< 0.001** | **< 0.001** | **< 0.001** | **< 0.001** |
| **BMI** | **< 0.001** | **< 0.001** | **< 0.001** | **< 0.001** | **< 0.001** | **< 0.001** | **< 0.001** | **< 0.001** | **< 0.001** | **< 0.001** | **< 0.001** | **< 0.001** | **< 0.001** |
| **Ethnicity** | **< 0.001** | **0.08** | **< 0.001** | **< 0.001** | **< 0.001** | **< 0.001** | **< 0.001** | **< 0.001** | **< 0.001** | **< 0.001** | **< 0.001** | **< 0.001** | **< 0.001** |
| **Education** | **< 0.001** | **< 0.001** | **< 0.001** | **< 0.001** | **< 0.001** | **< 0.001** | **< 0.001** | **< 0.001** | **< 0.001** | **< 0.001** | **< 0.001** | **< 0.001** | **< 0.001** |
| **Socio-economic status** | **< 0.001** | **< 0.001** | **< 0.001** | **< 0.001** | **< 0.001** | **< 0.001** | **< 0.001** | **< 0.001** | **< 0.001** | **< 0.001** | **< 0.001** | **< 0.001** | **< 0.001** |

Values are *p* values
